# Knowledge and motivations of training in peer review: an international cross-sectional survey

**DOI:** 10.1101/2022.09.03.22279564

**Authors:** Jessie V. Willis, Janina Ramos, Kelly D. Cobey, Jeremy Y. Ng, Hassan Khan, Marc A. Albert, Mohsen Alayche, David Moher

## Abstract

**Background:** Despite its globally accepted use in scholarly publishing, peer review is currently an unstandardized process lacking uniform guidelines. Previous surveys have demonstrated that peer reviewers, especially early career researchers, feel unprepared and undertrained to effectively conduct peer review. The purpose of this study was to conduct an international survey on the current perceptions and motivations of researchers regarding peer review training.

**Methods:** A cross-sectional online survey was conducted of biomedical researchers. Participants were identified using a random sample of 100 medical journals from a Scopus source list. A total of 2000 randomly selected corresponding authors from the last 20 published research articles from each journal were invited. The survey was administered via SurveyMonkey, participation in the survey was voluntary and all data was anonymized. An invite was sent via email on May 23 2022. Reminder emails were sent one and two weeks from the original invitation and the survey closed after three weeks. Participants were excluded from data analysis if less than 80% of questions were answered. Data was analyzed using SPSS Statistics and Microsoft Excel. Quantitative items were reported using frequencies and percentages or means and SE, as appropriate. A thematic content analysis was conducted for qualitative items in which two researchers independently assigned codes to the responses for each written-text question, and subsequently grouped the codes into themes. At both stages, conflicts were resolved through discussion until a consensus was achieved. A descriptive definition of each category was then created and unique themes – as well as the number and frequency of codes within each theme – were reported.

**Results:** A total of 186 participants completed the survey of the 2000 researchers invited. The average completion rate was 92% and it took on average 13 minutes to complete the survey. Fourteen responses were excluded based on having less than 80% questions answered. A total of 97 of 172 respondents (57.1%) identified as men. The majority (n = 108, 62.8%) were independent researchers defined as assistant, associate, or full professors of an academic organization (n = 103, 62.8%) with greater than 21 peer-reviewed articles published (n = 106, 61.6%). A total of 144 of 171 participants (84.2%) indicated they had never received formal training in peer review. Most participants (n = 128, 75.7%) agreed – of which 41 (32.0%) agreed strongly – that peer reviewers should receive formal training in peer review prior to acting as a peer reviewer. The most preferred training formats were all online, including online courses, lectures, and modules. A total of 55 of 80 (68.8%) participants indicated that their affiliated journal did not require peer review training for reviewers. In the thematic analysis of qualitative questions, the most common themes were related to providing clearer standards, expectations, and better incentives for reviewers. Most respondents (n = 111 of 147, 75.5%) stated that difficulty finding and/or accessing training was a barrier to completing training in peer review.

**Conclusion:** Despite being desired, most biomedical researchers have not received formal training in peer review and indicated that training was difficult to access or not available.

## INTRODUCTION

Peer review is the predominant quality control measure for scientific publishing regardless of country or discipline^1-3^. Peer review refers to the process by which “peers” are selected to assess the validity and quality of submitted manuscripts for publication^4^. Responsibilities of peer reviewers typically include providing constructive feedback to the authors of the manuscript and sometimes recommendations to journal editors^5,6^.

Despite its foothold in scholarly publishing, peer review is not a standardized process and lacks uniform guidelines^7-10^. Different scholarly publishers have different requirements and responsibilities for their peer reviewers and peer review data is not always made public^11^. Some publishers provide guidelines and training for their peer review process; however, a 2012 study found that only 35% of selected journals provided online instructions for their peer reviewers^12,13^.

It is therefore understandable that many potential peer reviewers feel inadequately trained to peer review. This is especially true for early career researchers; a recent survey showed that 60% of those under 36 years of age felt there is a lack of guidance on how to review papers^14^. Additional studies have shown that training is highly desired by academics^15-17^. In a 2018 survey by Publons, 88% of survey respondents felt training would have a positive impact on the efficacy of peer review. Despite this, 39% of respondents had never received training and 35.8% had self-trained by reading academic literature. Most respondents believed that training should be provided by scholarly publishers or journals and 45.5% believe that it should be a practical online course^18^.

Unfortunately, the effectiveness of peer review training has been studied only via small-scale studies on non-online methods (e.g., workshops) with limited evidence of any benefit^19-22^. Our group was unable to identify any randomized-controlled trials regarding how the electronic delivery of peer review guidelines has impacted the knowledge of potential peer reviewers.

In the present study we conducted a large-scale, online survey to provide an up-to-date perspective of international biomedical researchers’ views on peer review training. We focused on biomedical researchers as the needs and perspectives of researchers related to peer review may differ by discipline.

## METHODS

### Transparency Statement

Ethics approval was obtained from the Ottawa Health Science Network Research Ethics Board (OHSN-REB Protocol Number 20220237-01H). The study protocol was registered to the Open Science Framework (OSF) prior to data analysis (https://osf.io/wgxc2/)^23^. Text for this manuscript was drawn directly in reference to the registered protocol on OSF. Anonymous study data and any analytical code was shared publicly using the OSF and study findings were reported in a preprint and open access publication.

### Study Design

We conducted a cross-sectional online survey of biomedical researchers. The CHERRIES reporting guidelines were used to inform the reporting of our findings^24^.

### Participant sampling framework

We identified a random sample of international biomedical researchers who are actively publishing in peer-reviewed medical journals. We used the Scopus source list to randomly select 100 biomedical journals. The Scopus list was restricted to those journals with an All Science Journal Classification (ASJC) code of ‘Medicine’ and those that specified the journal was ‘active’ at the time of searching (November 2021). We excluded journals that indicated that they only published articles in a language other than English. Using the RAND function in Excel, we then randomly selected 100 journals from this list. Subsequently, we visited each of the randomly selected journal websites and extracted the corresponding authors from the last 20 published research articles. Corresponding author email extraction was completed on December 9, 2021. In instances where the journal was not open access and we did not have access via our academic institution, we replaced the journal with another randomly selected journal. We also replaced any journals which had non-functioning links. We have used this broad approach to sampling successfully in previous research^25^. This approach enabled us to identify a population of 2000 randomly selected researchers to invite to our survey.

### Survey

The survey was purposefully built for this study and was administered using SurveyMonkey software (https://www.surveymonkey.ca/r/7B2JYR6). This was a closed survey; thus, only available to invited participants via our sampling framework. We emailed the identified sample population a recruitment script with a link to the survey (Appendix 1). Participants were provided with a consent form prior to entering the survey and consent was presumed if they completed the survey. Participation in the survey was voluntary and all data was completely anonymized. The survey was sent on May 23, 2022. We sent all participants a reminder email to complete our survey after one week (May 30, 2022) and two weeks (June 6, 2022), respectively, from the original invitation. The survey was closed after three weeks (June 13, 2022).

The survey contained 37 questions: 1-10 were demographic questions about the participant, 11-15 were regarding level of experience with peer review, 16-23 were opinion-based questions about peer review training, 24-33 were for respondents who have experience running peer review from a journal perspective, and 34-37 were open-ended questions with comment boxes. 33 of the questions were quantitative while four were qualitative. The survey questions were presented in blocks based on content and question type. The survey used adaptive questioning where certain questions appeared based on the participants’ previous responses. The full list of survey questions can be found in Appendix 2.

The survey was created in SurveyMonkey by two authors (JVW, JR). All survey questions were reviewed and piloted by four researchers (HK, JYN, KDC, DM) and two invited researchers outside of the author list. The average time to complete the survey was estimated to be 15 minutes by pilot testers. All questions were optional and could be skipped. We offered participants the option to report their email to be entered into a draw to win one of three $100 Amazon Gift Cards. Email addresses were stored separately from the study data.

### Data Analysis

We used SPSS Statistics and Microsoft Excel for data analysis. We reported the overall response rate based on the number of individuals that completed our survey from the sample identified, as well as the survey completion rate (i.e., the number of people who viewed our survey that completed it). We excluded participants from data analysis if they did not complete 80% or more of the survey. We reported quantitative items using frequencies and percentages or means and SE, as appropriate. For qualitative items, we conducted a thematic content analysis of responses in Excel. For this, two researchers (JR, MAA) independently assigned codes to the responses for each written-text question. Codes were then discussed and iteratively updated until there was consensus among the two researchers that best reflected the data. Following this, individual codes were independently grouped into themes by the two reviewers and finalized by consensus. We then created a descriptive definition of each category. We reported the number of unique themes and the number and frequency of codes within each theme.

## RESULTS

### Protocol Amendments

Survey roll-out was changed from four weeks to three weeks due to time constraints. Minor revisions were made to the survey questions, recruitment and reminder emails and consent form.

### Participants

### Demographics

A total of 186 participants completed the survey of the 2000 researchers invited (9.3%). There were 107 (5.4%) instances where the email was unable to be sent and 32 (1.6%) instances where the participant indicated (including auto-replies) an inability to be reached/participate. As these accounted for less than 10% of invited participants, no changes were made to the recruitment strategy. A flowchart detailing these instances can be found in the supplementary material.

The average completion rate was 92% and it took, on average, 13 minutes to complete the survey. There were 14 responses that were excluded based on having less than 80% questions answered, thus the final included number was 172. A total of 97 respondents (57.1%) identified as men. The survey received responses from 48 different countries with the greatest representation from United States (n = 41, 24.0%), United Kingdom (n = 13, 7.6%) and India (n = 13, 7.6%). The majority (n = 108, 62.8%) were independent researchers defined as assistant, associate or full professors of an academic organization (n = 103, 62.8%) with greater than 21 peer-reviewed articles published (n = 106, 61.6%). Full demographics are described in Table 1.

**Table 1.** Demographic data.

|  |  | Frequency | Percent |
| --- | --- | --- | --- |
| Age | 18-24 | 1 | .6 |
|  | 25-34 | 30 | 17.4 |
|  | 35-44 | 60 | 34.9 |
|  | 45-54 | 35 | 20.3 |
|  | 55-64 | 26 | 15.1 |
|  | 65+ | 19 | 11.0 |
|  | Total | 171 | 99.4 |
| Gender | Man | 97 | 56.4 |
|  | Woman | 73 | 42.4 |
|  | Total | 170 | 98.8 |
| Occupation and/or Position | Other (please specify) | 22 | 12.8 |
|  | Master's student | 10 | 5.8 |
|  | PhD student | 12 | 7.0 |
|  | Post-doctoral fellow | 14 | 8.1 |
|  | Independent researcher (e.g., assistant/associate/full professor) | 108 | 62.8 |
|  | Research support staff (e.g., research assistant, research coordinator) | 6 | 3.5 |
|  | Total | 172 | 100.0 |
| Primary research interest | Other (please specify) | 58 | 33.7 |
|  | Clinical | 82 | 47.7 |
|  | Pre-clinical ("Basic science") | 30 | 17.4 |
|  | Total | 170 | 98.8 |
| Institution | Other (please specify) | 9 | 5.2 |
|  | University/college | 103 | 59.9 |
|  | Research institute | 4 | 2.3 |
|  | Healthcare institution (e.g., medical centre, hospital) | 42 | 24.4 |
|  | Private sector (e.g., pharmaceutical company) | 4 | 2.3 |
|  | Not-for-profit | 1 | .6 |
|  | Government organization | 7 | 4.1 |
|  | Total | 170 | 98.8 |
| Scholarly publishing experience | < 1 year | 1 | .6 |
|  | 1-5 years | 38 | 22.1 |
|  | 6-10 years | 44 | 25.6 |
|  | 11-15 years | 29 | 16.9 |
|  | 16-20 years | 13 | 7.6 |
|  | 21+ years | 47 | 27.3 |
|  | Total | 172 | 100.0 |
| Number of peer reviewed articles published to date | < 2 | 5 | 2.9 |
|  | 3-5 | 16 | 9.3 |
|  | 6-10 | 22 | 12.8 |
|  | 11-20 | 23 | 13.4 |
|  | 21-50 | 36 | 20.9 |
|  | 51+ | 70 | 40.7 |
|  | Total | 172 | 100.0 |

### Experience with peer review and peer review training

In total, 144 of 171 participants (84.2%) have never received formal training in peer review. The majority answered that their primary institution did not offer peer review training (n = 108, 63.2%) or otherwise did not know of any training offered (n = 48, 28.1%). For the 27 participants that had received peer review training, the most common training formats were in-person lectures (n = 12, 44.4%), online lectures (n = 10, 37.0%), or online courses of at least 6 sessions (n = 10, 37.0%). Most of the training received was provided by an academic organization (n = 18, 66.7%). Less than half (40.7%) of participants indicated the training was completed over 5 years ago.

For their first-time performing peer review, 88 of 166 (53.0%) participants felt either very unprepared (10.8%), unprepared (24.1%), or slightly unprepared (18.1%). Responses about peer review and peer review training are shown in Table 2.

**Table 2.** Experience with peer review and peer review training.

|  |  | Frequency | Percent |
| --- | --- | --- | --- |
| How many articles have your peer reviewed in the last 12 months? | 0 | 7 | 4.1 |
|  | 1-3 | 41 | 23.8 |
|  | 4-6 | 38 | 22.1 |
|  | 6-10 | 23 | 13.4 |
|  | >10 | 58 | 33.7 |
|  | I have never been a peer reviewer | 4 | 2.3 |
|  | Total | 171 | 99.4 |
| For how many years have you been active as a manuscript peer reviewer? | < 1 year | 11 | 6.4 |
|  | 1-5 years | 59 | 34.3 |
|  | 6-10 years | 43 | 25.0 |
|  | 11-15 years | 15 | 8.7 |
|  | 16-20 years | 13 | 7.6 |
|  | 21 + years | 28 | 16.3 |
|  | Total | 169 | 98.3 |
| Have you completed any formal training in peer review? | Yes | 26 | 15.1 |
|  | No | 144 | 83.7 |
|  | Unsure | 1 | .6 |
|  | Total | 171 | 99.4 |
| Does the primary institution you are affiliated with offer formal training in peer review? | Yes, and I have completed it | 10 | 5.8 |
|  | Yes, but I have not completed it | 5 | 2.9 |
|  | No | 108 | 62.8 |
|  | Unsure/don't know | 48 | 27.9 |
|  | Total | 171 | 99.4 |
| Type of formal peer review training completed |  | Frequency | Percent |
|  | Online lecture | 10 | 16.4 |
|  | Online course (at least 6 sessions) | 10 | 16.4 |
|  | In-person lecture | 12 | 19.7 |
|  | In-person half day workshop | 2 | 3.3 |
|  | In-person full day workshop | 7 | 11.5 |
|  | Shadowing a mentor/ghost-writing | 4 | 6.6 |
|  | Self-selected reading material | 7 | 11.5 |
|  | Online resource/module | 8 | 13.1 |
|  | Other | 1 | 1.6 |
|  | Total | 61 | 100.0 |
| Who provided peer review training that was completed | Journal | 4 | 12.1 |
|  | Publisher (of multiple journals) | 6 | 18.2 |
|  | University/college | 18 | 54.5 |
|  | Private company | 2 | 6.1 |
|  | Unsure/don't know | 2 | 6.1 |
|  | Other | 1 | 3.0 |
|  | Total | 33 | 100.0 |
| Peer review training provided by institution | Online lecture | 6 | 20.7 |
|  | Online course (at least 6 sessions) | 2 | 6.9 |
|  | In-person lecture | 3 | 10.3 |
|  | Half day workshop | 3 | 10.3 |
|  | Full day workshop | 5 | 17.2 |
|  | Shadowing a mentor/ghost-writing | 2 | 6.9 |
|  | Self-selected reading material | 2 | 6.9 |
|  | Online resource/modules | 4 | 13.8 |
|  | Unsure/don't know | 1 | 3.4 |
|  | Other | 1 | 3.4 |
|  | Total | 29 | 100.0 |

### Opinion-based Questions

#### General statements on peer review and peer review training

Participants rated their agreement with statements related to peer review and peer review training on a 7-point scale from strongly disagree to strongly agree. A graph of the responses is depicted in Figure 1.

**Figure 1.**
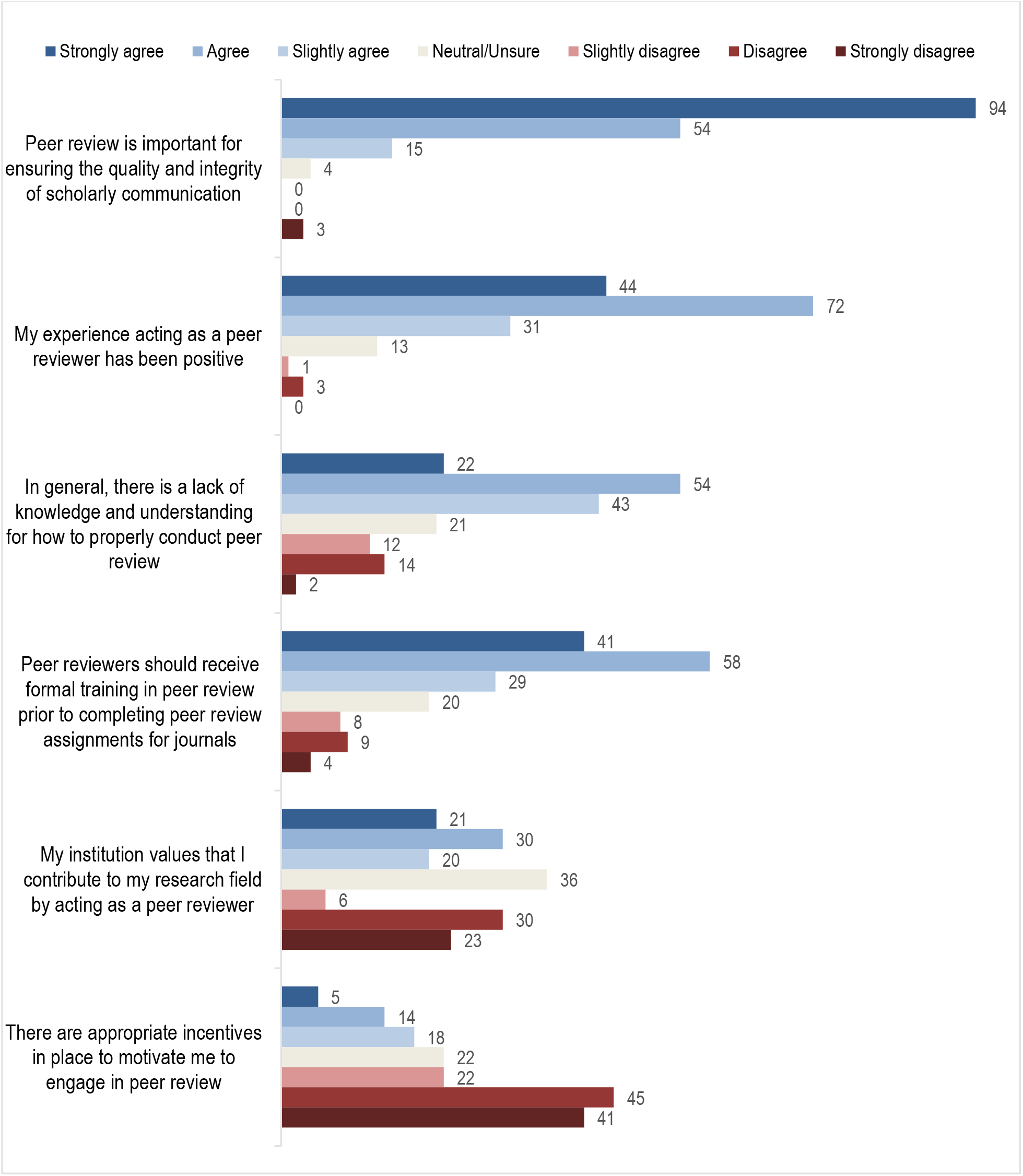
Participant agreement with statements based on overall experiences with peer review in the last 12 months.

Notable findings included that 148 respondents (86.5%) either strongly agreed or agreed that peer review is important for ensuring the quality and integrity of scholarly communication. One hundred and sixteen (69.5%) strongly agreed or agreed that their experience as a peer reviewer had been positive. Seventy-six (45.2%) strongly agreed or agreed that there is a lack of knowledge and understanding for how to properly conduct peer review. Ninety-nine (58.9%) strongly agreed or agreed that peer reviewers should receive formal training in peer review prior to acting as a peer reviewer for journals. Eighty-six (50.9%) strongly disagreed or disagreed that there were appropriate incentives in place to motivate them to engage in peer review.

#### Desired training topics, organizations and formats

These questions required participants to rank their responses in order from most to least preferred. A graph of the ranked responses can be found in Figure 2.

**Figure 2.**
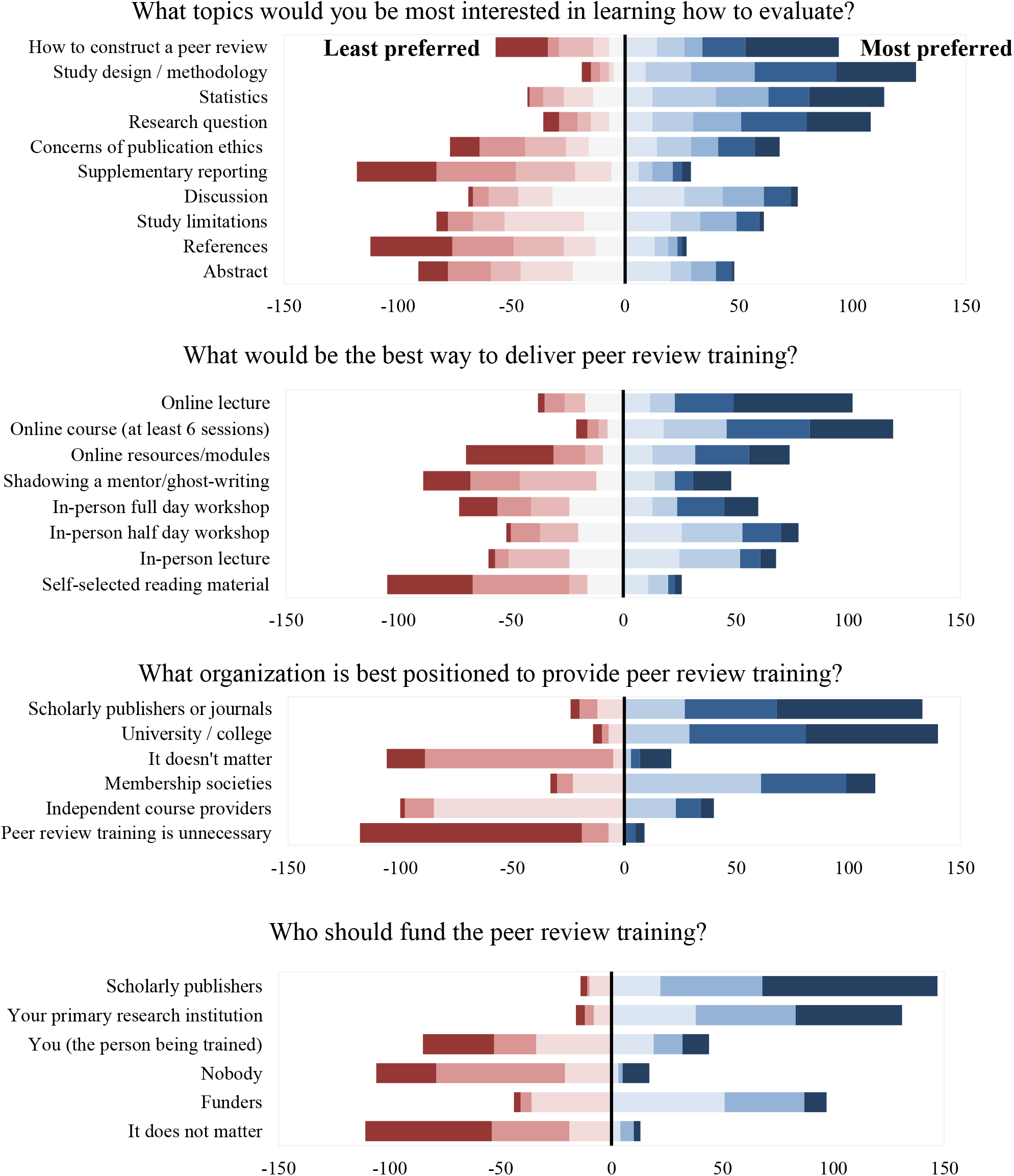
Ranking of preferred topics, training format, and creating organizations. Y-axis in order by number of times ranked number one (least to most, bottom to top).

The topic that was most frequently ranked as the most desired was how to construct a peer review. Based on average rank placement, the most desired topics were appraisal of study design and methodology, appraisal of the research question and appraisal of statistics. The most desired training formats were all online, including online lectures, online courses (at least 6 sessions), and online resources or modules.

For the organization type to deliver the training, scholarly publishers or journals were ranked as most preferred more often than academic organizations. However, based on average rank placement, academic organizations were most preferred overall. The most preferred funding organization was scholarly publishers, followed by the primary research institution of the trainee.

#### Journal-specific questions

Participants were only able to answer these questions if they indicated they worked or volunteered for a journal that publishes peer reviewed articles. There were 80 respondents that were included in this section.

In total, 55 of 80 participants (68.8%) indicated that the journal they were affiliated with did not explicitly require peer review training for their reviewers. Eight (10.0%) indicated that it was required and provided by the journal internally, while two (2.5%) indicated that it was required by externally delivered. The most common format of training required was either an online course and/or lecture. Required training length was variable from 1-5 hours to 20+ hours.

Only 10 of 80 (10.0%) of participants indicated that the journal assessed peer review reports of new reviewers; however, the majority (n = 51, 63.8%) indicated they were unsure or did not know. Twenty-one (26.6%) provided reporting guidelines to reviewers as part of the peer review assessment process.

#### Qualitative questions

The full thematic content analysis can be found in the supplementary files. The most covered themes for each qualitative question are shown in Table 3.

**Table 3.** Thematic content analysis for qualitative questions.

|  |  |
| --- | --- |
| ***Other than training, how do you believe the quality of peer review could be improved?*** | **152** |
| Clear standards and expectations for peer reviewers | 36 |
| Improved incentives | 32 |
| Increased feedback and oversight | 28 |
| ***What barriers do you face in engaging in peer review training?*** | **147** |
| Difficulty finding and accessing training | 111 |
| Lack of personal incentive | 15 |
| Complexity and inconsistency of peer review requirements | 8 |
| Lack of value placed on peer review or training | 8 |
| ***What would incentivize you to obtain additional training in peer review best practices?*** | **136** |
| Recognition for training or peer review | 39 |
| Financial incentives | 35 |
| Clear translation into the peer review process | 27 |
| ***Any other comments you wish to share relating to peer review training:*** | **68** |
| Call for improvement of the peer review system | 17 |
| Recommendation regarding providing training | 16 |
| Related to incentives | 10 |

## DISCUSSION

Our international survey demonstrated that most biomedical researchers completing peer review have received no formal training, although most agreed that training should be a requirement.

Other professional competences typically require training and certification. For example, a physician in North America requires over four years of training, evaluation, and certification to practice medicine. Comparatively, peer review does not require any training, but is a crucial activity in the publication of the very same evidence-based medicine used by physicians. For example, poor quality publications could be referenced in systematic reviews which may in turn inform clinical guidelines, thus having an impact on the general population.

A lack of training could therefore help to explain the less-than-optimal reporting quality of biomedical research^26^ and the inability of reviewers to detect major errors and deficiencies^27^. A recent systematic review conducted by our team demonstrated the limited availability of openly accessible online peer review training^28^. As indicated by our survey, most respondents reported that a large barrier to receiving training was the limited availability and accessibility of training material.

We feel that it is important to address the current shortage of openly accessible peer review training as indicated by our survey and recent systematic review. Using the results of our two studies, we will map out a potential path forward to enhance a degree of professionalism in peer review.

First, there needs to be agreement across players (e.g., researchers; editors; peer reviewers) regarding a core set of competences that are minimally necessary for all peer reviewers. This has previously been done for editors^29^. Our recent systematic review of online training opportunities in peer review indicated that most available training focused on process issues, such as how to effectively write a peer review report. Less attention was paid to the use of reporting guidelines or the critical appraisal of statistics or clinical trials^28^. As indicated in our survey, the most preferred topics were critical appraisal of methodology and statistical analysis.

Second, the development of educational and training modules is required to flesh out these competencies as a minimum set of evidence-based pedagogical sessions. Our recent systematic review demonstrated that most available training could be completed in less than an hour, which may indicate that current training is not comprehensive^28^. As indicated by our survey, both online lectures and online courses of at least six sessions are highly desired.

Third, ensuring that training is openly accessible to learners is in keeping with equity, diversity, and inclusiveness principles. In our recent systematic review, most online training was not openly accessible and had a barrier to access (e.g., membership requirements, paywall). To maximize dissemination and uptake, a Massive Open Online Course (MOOC) needs to be developed for delivery in multiple languages. Recent studies have shown the ability of MOOCs to reach a diverse audience in healthcare^30,31^. Fourth is the need to evaluate participants. Those who are successful at meeting an agreed-upon threshold should be credentialed as certified peer reviewers. Fifth and final, is a need to evaluate the effectiveness of the training program.

Furthermore, the target audience of the training should be discussed. Another study demonstrated that early career researchers (ECRs) desire greater guidance in peer review^14^. It is important to note the careful balance between enriching the potential peer reviewer pool and further disincentivizing peer review, as journals currently face challenges finding peer reviewers, in part due the sequala of COVID-19^18^. In our survey, some respondents stated that they are not personally motivated to obtain formal training in peer review given their experience level. Additionally, requiring training for all peer reviewers may also increase the time to publication. We would argue, however, that training early career researchers with little to no experience in peer review could help address the shortage of people willing to peer review.

Currently, journal editors may spend time emailing numerous potentially untrained peer reviewers when they could instead contact a smaller number of highly trained and motivated peer reviewers. Additionally, increasing incentives for peer review was mentioned by respondents of our survey and additionally has been the focus of other studies in peer review^32,33^.

Limitations to our study include the overall experience level of the respondents. Most respondents to our survey were well established researchers, as we pulled from a list of corresponding authors on published manuscripts. Additionally, our survey had a response rate of only 9.3%. Therefore, whether non-responders and early career researchers would respond similarly or report a greater need for training is unknown.

Lastly, our study results indicate a strong view that training in peer review should be offered by institutions and/or publishers. The latter, particularly the publishing oligopoly, have the fiscal resources to enable peer review training. The former might have the scholars with the expertise to deliver comprehensive training but likely do not have the resources to deliver the training. Currently, peer review is largely an activity without fiscal cost to editors and publishers. Stakeholders in peer review should focus future efforts on creating an openly accessible training program in peer review. We ask that journals indicate their support for training in manuscript peer review, their interest in engaging in the development of a core competency program, and their willingness to participate in the roll out and evaluation of a training program.

## Supporting information

Thematic content analysis

Survey responses_SPSS

Survey responses_raw

Email flowchart

## Data Availability

All data produced are available online at Open Science Framework.

https://osf.io/wgxc2/

## APPENDICES Appendix 1

Recruitment script

### Initial Recruitment Email

Subject Line: Invitation to Participate in Peer Review Training Survey

Dear <NAME>,

I hope this email finds you well. My name is Janina Ramos and I am emailing on behalf of Dr. David Moher at the Centre for Journalology, Ottawa Hospital Research Institute. We obtained a random sample of corresponding authors of recently published articles in biomedical journals. Your name was among those captured in our sample and we are writing to invite you to contribute to a short survey about peer review training.

The study has received ethical approval through the Ottawa Hospital Health Research Network’s Research Ethics Board (OHSN-REB Protocol Number 20220237-01H) and you will be provided with an online consent form. By completing the survey, you are providing your consent to participate in the study. Your participation is voluntary. Your responses will be completely anonymized. We anticipate the survey taking approximately 15 minutes.

This study will be the first to provide a current international perspective on biomedical researchers’ knowledge and perceptions of, and engagement with, peer review training. Your participation in this survey will help us to help identify gaps in peer review training experience and knowledge. Subsequently, it may guide the creation of future training options, inform the development of preferred training methods, and increase comprehensiveness of peer review training for biomedical researchers.

#### If you would like to participate in this study, please click on the link below to view the consent form and access the survey: https://www.surveymonkey.ca/r/7HXYQYN

Should you have any questions or concerns about our invitation, please do not hesitate to reach out to our principal investigator, Dr. David Moher, PhD at.

Thank you,

Janina Ramos (on behalf of the research team)

### Reminder Email

Subject Line: Reminder: Invitation to Participate in Peer Review Training Survey Dear <NAME>,

We hope this email finds you well. This email serves as a gentle reminder of our request to participate in our research study sent out <time period> weeks ago. We thank you for your time, if you have already participated!

My name is Janina Ramos and I am emailing on behalf of Dr. David Moher at the Centre for Journalology, Ottawa Hospital Research Institute. We obtained a random sample of corresponding authors of recently published articles in biomedical journals. Your name was among those captured in our sample and we are writing to invite you to contribute to a short survey about peer review training.

The study has been reviewed by the Ottawa Hospital Health Research Network’s Research Ethics Board (OHSN-REB Protocol Number 20220237-01H. and you will be provided with an online consent form. By completing the survey, you are providing your consent to participate in the study. Your participation is voluntary. Your responses will be completely anonymized. We anticipate the survey taking approximately 15 minutes.

This study will be the first to provide a current international perspective on biomedical researchers’ knowledge and perceptions of, and engagement with, peer review training. Your participation in this survey will help us to help identify gaps in peer review training experience and knowledge. Subsequently, it may guide the creation of future training options, inform the development of preferred training methods, and increase comprehensiveness of peer review training for biomedical researchers.

#### If you would like to participate in this study, please click on the link below to view the consent form and access the survey: https://www.surveymonkey.ca/r/7HXYQYN

We will send all participants a reminder email to complete our survey after 1 and 2 weeks from the original invitation, the survey will close after 3 weeks. Should you have any questions or concerns about our invitation, please do not hesitate to reach to our principal investigator, Dr. David Moher, PhD at.

Thank you,

Janina Ramos (on behalf of the research team)

## Appendix 2

Survey items

### Demographics

| # | Question | Answer |
| --- | --- | --- |
| 1 | What is your age group? | a) 18-24<br>b) 25-34<br>c) 35-44<br>d) 45-54<br>e) 55-64<br>f) 65 + |
| 2 | What is your gender identity? | a) Man<br>b) Woman<br>c) Non-binary<br>d) Prefer to self-describe, below:<br>e) Prefer not to say |
| 3 | What country do you live in? | [drop down list] |
| 4 | Which describes you best? | a) Undergraduate student<br>b) Master's student<br>c) PhD student<br>d) Post-doctoral fellow<br>e) Independent researcher (e.g., assistant/associate/full professor)<br>f) Research support staff (e.g., research assistant, research coordinator)<br>g) Other, please specify: |
| 5 | How many years of experience do you have with scholarly publishing (i.e. writing and publishing manuscripts)? | a) < 1 year<br>b) 1-5 years<br>c) 6-10 years<br>d) 11-15 years<br>e) 16-20 years<br>f) 21 + years |
| 6 | How would you primarily describe the research you conduct? | a) Clinical<br>b) Pre-clinical ("basic" science)<br>c) Other, please specify: |
| 7 | How would you describe the institution of your primary occupation? | a) University/college<br>b) Research institute<br>c) Healthcare institution (e.g., medical centre, hospital)<br>d) Private sector (e.g., pharmaceutical company)<br>e) Not-for-profit<br>f) Government organization<br>g) Other, please specify: |
| 8 | How many articles have you peer reviewed in the last 12 months? | a) 0<br>b) 1-3<br>c) 4-6<br>d) 6-10<br>e) >10<br>f) I have never been a peer reviewer |
| 9 | For how many years have you been active as a manuscript peer reviewer? | a) < 1 year<br>b) 1-5 years<br>c) 6-10 years<br>d) 11-15 years<br>e) 16-20 years<br>f) 21 + years |
| 10 | How many peer reviewed articles have you published to date? | a) < 2<br>b) 3-5<br>c) 6-10<br>d) 11-20<br>e) 21-50<br>f) 51+ |

### Questions relating to experience with peer review

| # | Question | Answer |
| --- | --- | --- |
| 11 | Have you completed any formal training in manuscript peer review? | a) Yes → go to question 12<br>b) No → go to question 15<br>c) Unsure → go to question 15 |
| 12 | What format of training best describes the training you received? Check all that apply. | a) Online lecture<br>b) Online course (at least 6 sessions)<br>c) In-person lecture<br>d) In-person half day workshop<br>e) In-person full day workshop<br>f) Shadowing a mentor/ghost-writing<br>g) Self-selected reading material<br>h) Online resources/modules<br>i) Other, please specify |
| 13 | Who provided the training you received? Check all that apply. | a) A journal<br>b) A publisher (of multiple journals)<br>c) A university/college<br>d) Private company<br>e) Unsure/don't know<br>f) Other, please specify: |
| 14 | When did you receive the training? | a) $\leq 1$ year ago<br>b) 2 years ago<br>c) 3 years ago<br>d) 4 years ago<br>e) $\geq 5$ years ago |
| 15 | Does the primary institution you are affiliated with offer formal training for peer review? | a) Yes and I have completed it → go to question 16<br>b) Yes but I have not completed it → go to question 16<br>c) No → go to next section<br>d) Unsure/don't know → go to next section |
| 16 | What type of training does your primary institution offer? Check all that apply. | a) Online lecture<br>b) Online course (at least 6 sessions)<br>c) In-person lecture<br>d) In-person half day workshop<br>e) In-person full day workshop<br>f) Shadowing a mentor/ghost-writing<br>g) Self-selected reading material<br>h) Online resources/modules<br>i) Unsure/don't know<br>j) Other, please specify |

### Opinion-based questions about manuscript peer review

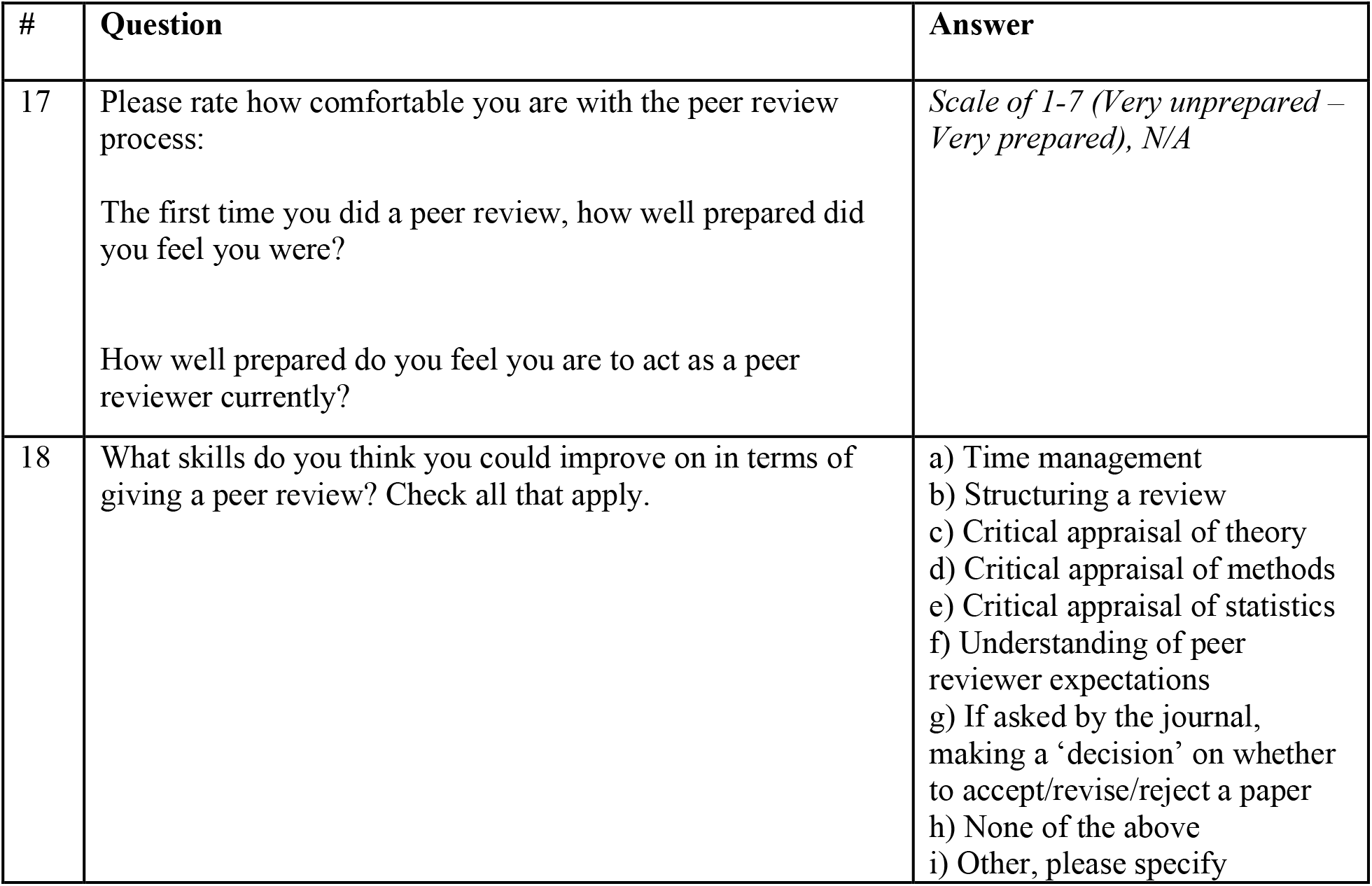
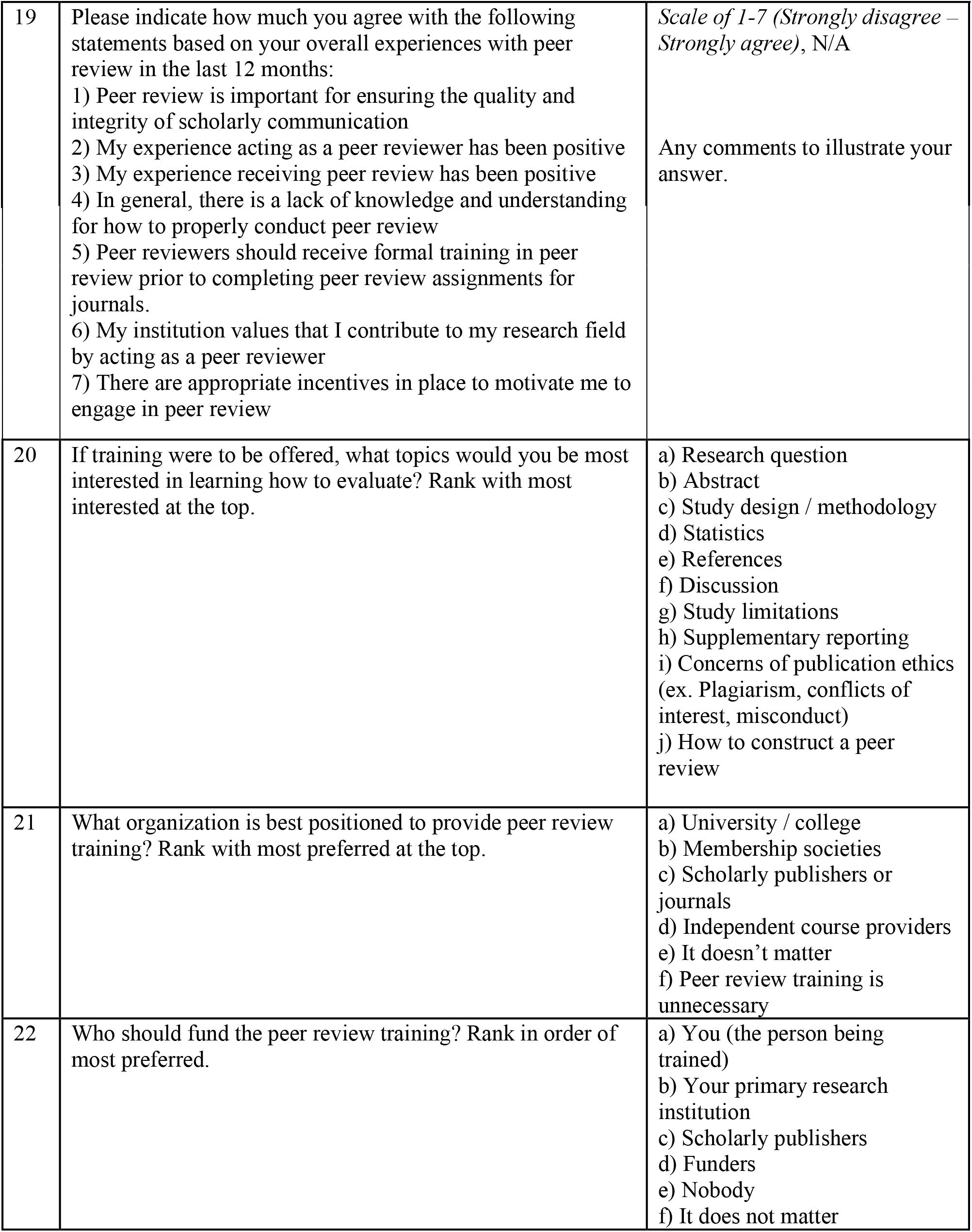
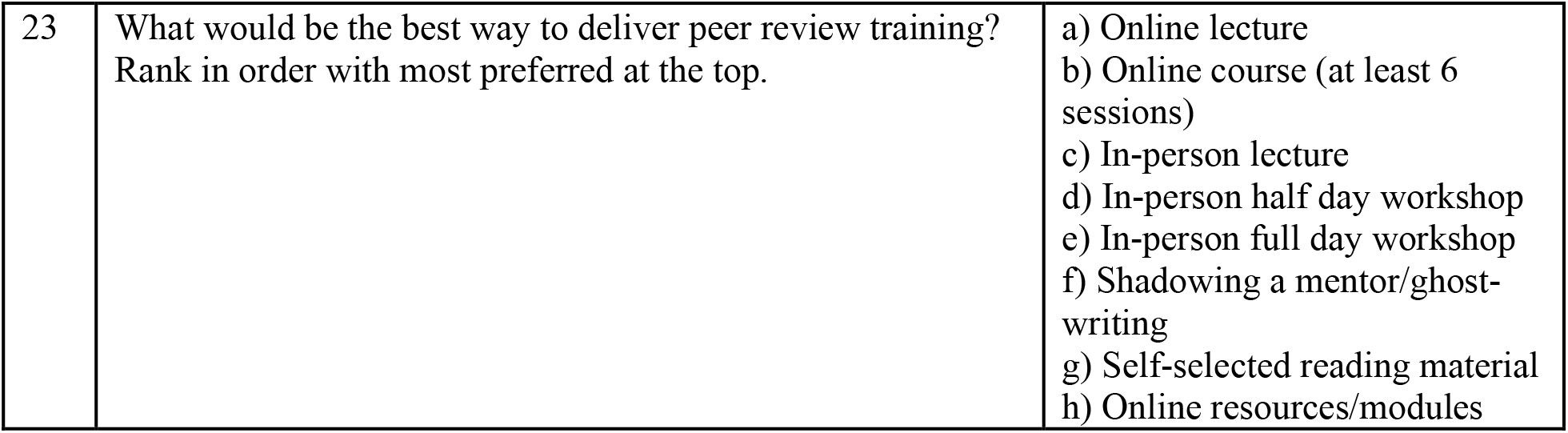

| # | Question | Answer |
| --- | --- | --- |
| 17 | Please rate how comfortable you are with the peer review process:<br>The first time you did a peer review, how well prepared did you feel you were?<br>How well prepared do you feel you are to act as a peer reviewer currently? | *Scale of 1-7 (Very unprepared – Very prepared), N/A* |
| 18 | What skills do you think you could improve on in terms of giving a peer review? Check all that apply. | a) Time management<br>b) Structuring a review<br>c) Critical appraisal of theory<br>d) Critical appraisal of methods<br>e) Critical appraisal of statistics<br>f) Understanding of peer reviewer expectations<br>g) If asked by the journal, making a 'decision' on whether to accept/revise/reject a paper<br>h) None of the above<br>i) Other, please specify |
| 19 | Please indicate how much you agree with the following statements based on your overall experiences with peer review in the last 12 months:<br>1) Peer review is important for ensuring the quality and integrity of scholarly communication<br>2) My experience acting as a peer reviewer has been positive<br>3) My experience receiving peer review has been positive<br>4) In general, there is a lack of knowledge and understanding for how to properly conduct peer review<br>5) Peer reviewers should receive formal training in peer review prior to completing peer review assignments for journals.<br>6) My institution values that I contribute to my research field by acting as a peer reviewer<br>7) There are appropriate incentives in place to motivate me to engage in peer review | *Scale of 1-7 (Strongly disagree – Strongly agree), N/A*<br>Any comments to illustrate your answer. |
| 20 | If training were to be offered, what topics would you be most interested in learning how to evaluate? Rank with most interested at the top. | a) Research question<br>b) Abstract<br>c) Study design / methodology<br>d) Statistics<br>e) References<br>f) Discussion<br>g) Study limitations<br>h) Supplementary reporting<br>i) Concerns of publication ethics (ex. Plagiarism, conflicts of interest, misconduct)<br>j) How to construct a peer review |
| 21 | What organization is best positioned to provide peer review training? Rank with most preferred at the top. | a) University / college<br>b) Membership societies<br>c) Scholarly publishers or journals<br>d) Independent course providers<br>e) It doesn't matter<br>f) Peer review training is unnecessary |
| 22 | Who should fund the peer review training? Rank in order of most preferred. | a) You (the person being trained)<br>b) Your primary research institution<br>c) Scholarly publishers<br>d) Funders<br>e) Nobody<br>f) It does not matter |
| 23 | What would be the best way to deliver peer review training?<br>Rank in order with most preferred at the top. | a) Online lecture<br>b) Online course (at least 6 sessions)<br>c) In-person lecture<br>d) In-person half day workshop<br>e) In-person full day workshop<br>f) Shadowing a mentor/ghost-writing<br>g) Self-selected reading material<br>h) Online resources/modules |

### Journal perspective

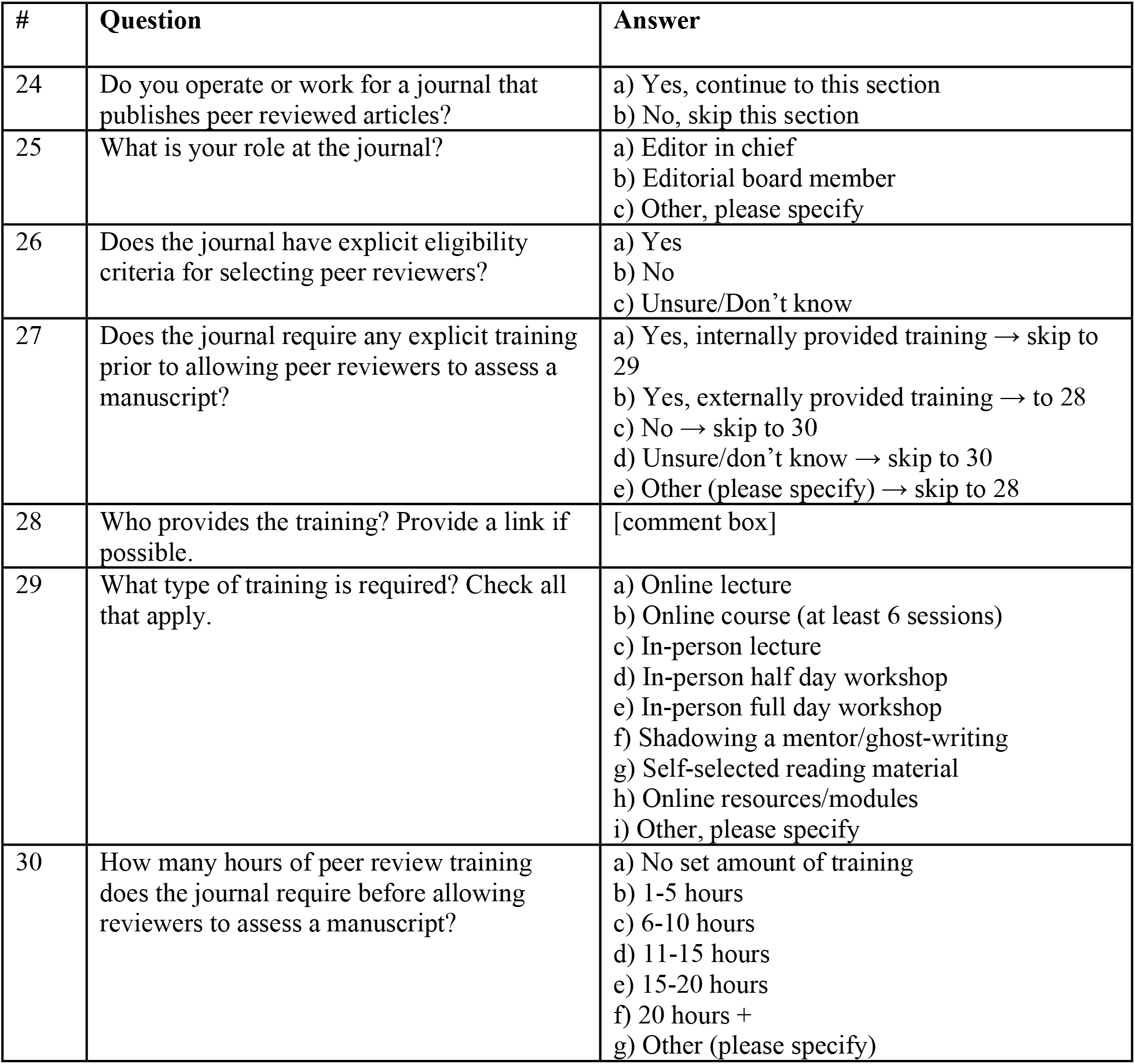
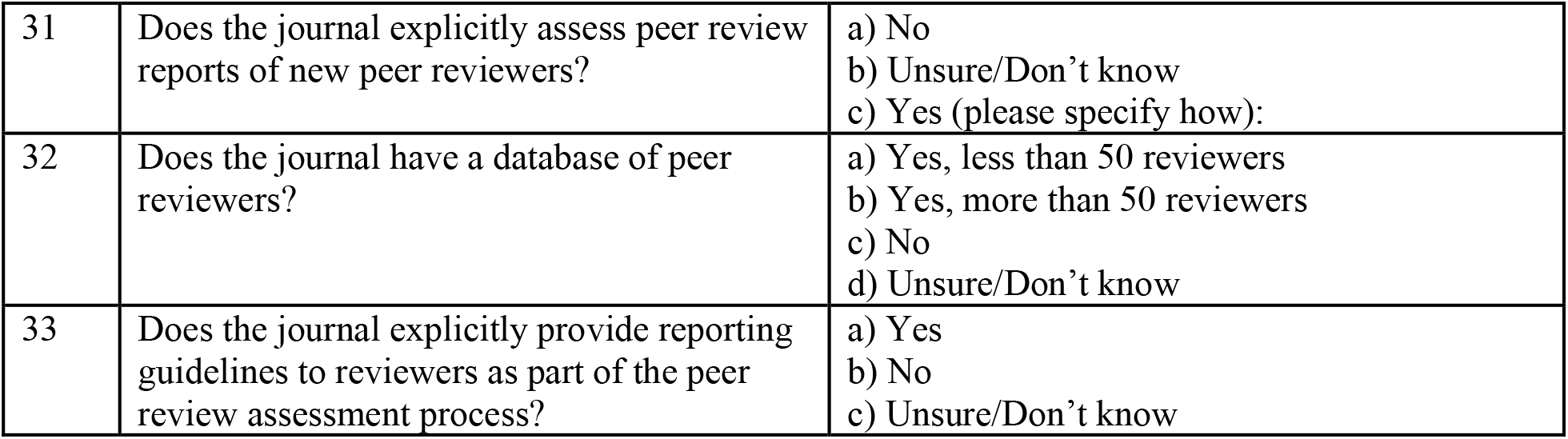

| # | Question | Answer |
| --- | --- | --- |
| 24 | Do you operate or work for a journal that publishes peer reviewed articles? | a) Yes, continue to this section<br>b) No, skip this section |
| 25 | What is your role at the journal? | a) Editor in chief<br>b) Editorial board member<br>c) Other, please specify |
| 26 | Does the journal have explicit eligibility criteria for selecting peer reviewers? | a) Yes<br>b) No<br>c) Unsure/Don't know |
| 27 | Does the journal require any explicit training prior to allowing peer reviewers to assess a manuscript? | a) Yes, internally provided training → skip to 29<br>b) Yes, externally provided training → to 28<br>c) No → skip to 30<br>d) Unsure/don't know → skip to 30<br>e) Other (please specify) → skip to 28 |
| 28 | Who provides the training? Provide a link if possible. | [comment box] |
| 29 | What type of training is required? Check all that apply. | a) Online lecture<br>b) Online course (at least 6 sessions)<br>c) In-person lecture<br>d) In-person half day workshop<br>e) In-person full day workshop<br>f) Shadowing a mentor/ghost-writing<br>g) Self-selected reading material<br>h) Online resources/modules<br>i) Other, please specify |
| 30 | How many hours of peer review training does the journal require before allowing reviewers to assess a manuscript? | a) No set amount of training<br>b) 1-5 hours<br>c) 6-10 hours<br>d) 11-15 hours<br>e) 15-20 hours<br>f) 20 hours +<br>g) Other (please specify) |
| 31 | Does the journal explicitly assess peer review reports of new peer reviewers? | a) No<br>b) Unsure/Don't know<br>c) Yes (please specify how): |
| 32 | Does the journal have a database of peer reviewers? | a) Yes, less than 50 reviewers<br>b) Yes, more than 50 reviewers<br>c) No<br>d) Unsure/Don't know |
| 33 | Does the journal explicitly provide reporting guidelines to reviewers as part of the peer review assessment process? | a) Yes<br>b) No<br>c) Unsure/Don't know |

### Open-ended

1. Other than training, how do you believe the quality of peer review could be improved?
2. What barriers do you face in engaging in peer review training?
3. What would incentivize you to obtain additional training in peer review best practices?
4. Any other comments you wish to share relating to peer review training:

## Notes

**Conflict of Interest** The authors have no conflicts of interest to declare.

### Competing Interest Statement

The authors have declared no competing interest.

### Clinical Protocols

https://osf.io/wgxc2/

### Funding Statement

This study did not receive any funding.

### Author Declarations

Ottawa Health Science Network Research Ethics Board of the Ottawa Hospital Research Institute gave ethical approval for this work.

