## Supplementary material for "Knowledge and motivations of training in peer review: an international cross-sectional survey": Survey responses_SPSS

| **What is your age group?** | | | | | |
| --- | --- | --- | --- | --- | --- |
|  | | Frequency | Percent | Valid Percent | Cumulative Percent |
| Valid | 18-24 | 1 | .6 | .6 | .6 |
|  | 25-34 | 30 | 17.4 | 17.5 | 18.1 |
|  | 35-44 | 60 | 34.9 | 35.1 | 53.2 |
|  | 45-54 | 35 | 20.3 | 20.5 | 73.7 |
|  | 55-64 | 26 | 15.1 | 15.2 | 88.9 |
|  | 65+ | 19 | 11.0 | 11.1 | 100.0 |
|  | Total | 171 | 99.4 | 100.0 |  |
| Missing | System | 1 | .6 |  |  |
| Total | | 172 | 100.0 |  |  |

| **What is your gender identity?** | | | | | |
| --- | --- | --- | --- | --- | --- |
|  | | Frequency | Percent | Valid Percent | Cumulative Percent |
| Valid | Man | 97 | 56.4 | 57.1 | 57.1 |
|  | Woman | 73 | 42.4 | 42.9 | 100.0 |
|  | Total | 170 | 98.8 | 100.0 |  |
| Missing | System | 2 | 1.2 |  |  |
| Total | | 172 | 100.0 |  |  |

| **What country do you live in?** | | | | | |
| --- | --- | --- | --- | --- | --- |
|  | | Frequency | Percent | Valid Percent | Cumulative Percent |
| Valid | United States | 41 | 23.8 | 24.0 | 24.0 |
|  | United Kingdom | 13 | 7.6 | 7.6 | 31.6 |
|  | Canada | 11 | 6.4 | 6.4 | 38.0 |
|  | Australia | 4 | 2.3 | 2.3 | 40.4 |
|  | Argentina | 2 | 1.2 | 1.2 | 41.5 |
|  | Bangladesh | 1 | .6 | .6 | 42.1 |
|  | Brazil | 4 | 2.3 | 2.3 | 44.4 |
|  | Switzerland | 3 | 1.7 | 1.8 | 46.2 |
|  | Germany | 1 | .6 | .6 | 46.8 |
|  | Denmark | 1 | .6 | .6 | 47.4 |
|  | Egypt | 2 | 1.2 | 1.2 | 48.5 |
|  | Spain | 1 | .6 | .6 | 49.1 |
|  | Ethiopia | 3 | 1.7 | 1.8 | 50.9 |
|  | France | 2 | 1.2 | 1.2 | 52.0 |
|  | Croatia | 3 | 1.7 | 1.8 | 53.8 |
|  | Indonesia | 1 | .6 | .6 | 54.4 |
|  | India | 13 | 7.6 | 7.6 | 62.0 |
|  | Iraq | 3 | 1.7 | 1.8 | 63.7 |
|  | Iran | 3 | 1.7 | 1.8 | 65.5 |
|  | Italy | 7 | 4.1 | 4.1 | 69.6 |
|  | Japan | 2 | 1.2 | 1.2 | 70.8 |
|  | Kenya | 1 | .6 | .6 | 71.3 |
|  | Lebanon | 1 | .6 | .6 | 71.9 |
|  | Mexico | 3 | 1.7 | 1.8 | 73.7 |
|  | Nigeria | 2 | 1.2 | 1.2 | 74.9 |
|  | Netherlands | 2 | 1.2 | 1.2 | 76.0 |
|  | Nepal | 4 | 2.3 | 2.3 | 78.4 |
|  | New Zealand | 1 | .6 | .6 | 78.9 |
|  | Philippines | 5 | 2.9 | 2.9 | 81.9 |
|  | Pakistan | 2 | 1.2 | 1.2 | 83.0 |
|  | Poland | 1 | .6 | .6 | 83.6 |
|  | West Bank | 1 | .6 | .6 | 84.2 |
|  | Portugal | 2 | 1.2 | 1.2 | 85.4 |
|  | Romania | 1 | .6 | .6 | 86.0 |
|  | Rwanda | 3 | 1.7 | 1.8 | 87.7 |
|  | Saudi Arabia | 2 | 1.2 | 1.2 | 88.9 |
|  | Sudan | 1 | .6 | .6 | 89.5 |
|  | Sweden | 2 | 1.2 | 1.2 | 90.6 |
|  | Singapore | 2 | 1.2 | 1.2 | 91.8 |
|  | Slovenia | 1 | .6 | .6 | 92.4 |
|  | Turks and Caicos Islands | 1 | .6 | .6 | 93.0 |
|  | Thailand | 1 | .6 | .6 | 93.6 |
|  | Tunisia | 3 | 1.7 | 1.8 | 95.3 |
|  | Turkey | 1 | .6 | .6 | 95.9 |
|  | Ukraine | 1 | .6 | .6 | 96.5 |
|  | Uganda | 1 | .6 | .6 | 97.1 |
|  | South Africa | 4 | 2.3 | 2.3 | 99.4 |
|  | Zimbabwe | 1 | .6 | .6 | 100.0 |
|  | Total | 171 | 99.4 | 100.0 |  |
| Missing | System | 1 | .6 |  |  |
| Total | | 172 | 100.0 |  |  |

| **Which describes you best? (Occupation/Position)** | | | | | |
| --- | --- | --- | --- | --- | --- |
|  | | Frequency | Percent | Valid Percent | Cumulative Percent |
| Valid | Other (please specify) | 22 | 12.8 | 12.8 | 12.8 |
|  | Master's student | 10 | 5.8 | 5.8 | 18.6 |
|  | PhD student | 12 | 7.0 | 7.0 | 25.6 |
|  | Post-doctoral fellow | 14 | 8.1 | 8.1 | 33.7 |
|  | Independent researcher (e.g. assistant/associate/full professor) | 108 | 62.8 | 62.8 | 96.5 |
|  | Research support staff (e.g. research assistant, research coordinator) | 6 | 3.5 | 3.5 | 100.0 |
|  | Total | 172 | 100.0 | 100.0 |  |

| **How many years of experience do you have with scholarly publishing (i.e. writing and publishing manuscripts)?** | | | | | |
| --- | --- | --- | --- | --- | --- |
|  | | Frequency | Percent | Valid Percent | Cumulative Percent |
| Valid | < 1 year | 1 | .6 | .6 | .6 |
|  | 1-5 years | 38 | 22.1 | 22.1 | 22.7 |
|  | 6-10 years | 44 | 25.6 | 25.6 | 48.3 |
|  | 11-15 years | 29 | 16.9 | 16.9 | 65.1 |
|  | 16-20 years | 13 | 7.6 | 7.6 | 72.7 |
|  | 21+ years | 47 | 27.3 | 27.3 | 100.0 |
|  | Total | 172 | 100.0 | 100.0 |  |

| **How would you primarily describe the research you conduct?** | | | | | |
| --- | --- | --- | --- | --- | --- |
|  | | Frequency | Percent | Valid Percent | Cumulative Percent |
| Valid | Other (please specify) | 58 | 33.7 | 34.1 | 34.1 |
|  | Clinical | 82 | 47.7 | 48.2 | 82.4 |
|  | Pre-clinical ("Basic science") | 30 | 17.4 | 17.6 | 100.0 |
|  | Total | 170 | 98.8 | 100.0 |  |
| Missing | System | 2 | 1.2 |  |  |
| Total | | 172 | 100.0 |  |  |

| **How would you describe the institution of your primary occupation?** | | | | | |
| --- | --- | --- | --- | --- | --- |
|  | | Frequency | Percent | Valid Percent | Cumulative Percent |
| Valid | Other (please specify) | 9 | 5.2 | 5.3 | 5.3 |
|  | University/college | 103 | 59.9 | 60.6 | 65.9 |
|  | Research institute | 4 | 2.3 | 2.4 | 68.2 |
|  | Healthcare institution (e.g. medical centre, hospital) | 42 | 24.4 | 24.7 | 92.9 |
|  | Private sector (e.g. pharmaceutical company) | 4 | 2.3 | 2.4 | 95.3 |
|  | Not-for-profit | 1 | .6 | .6 | 95.9 |
|  | Government organization | 7 | 4.1 | 4.1 | 100.0 |
|  | Total | 170 | 98.8 | 100.0 |  |
| Missing | System | 2 | 1.2 |  |  |
| Total | | 172 | 100.0 |  |  |

| **How many articles have you peer reviewed in the last 12 months?** | | | | | |
| --- | --- | --- | --- | --- | --- |
|  | | Frequency | Percent | Valid Percent | Cumulative Percent |
| Valid | 0 | 7 | 4.1 | 4.1 | 4.1 |
|  | 1-3 | 41 | 23.8 | 24.0 | 28.1 |
|  | 4-6 | 38 | 22.1 | 22.2 | 50.3 |
|  | 6-10 | 23 | 13.4 | 13.5 | 63.7 |
|  | >10 | 58 | 33.7 | 33.9 | 97.7 |
|  | I have never been a peer reviewer | 4 | 2.3 | 2.3 | 100.0 |
|  | Total | 171 | 99.4 | 100.0 |  |
| Missing | System | 1 | .6 |  |  |
| Total | | 172 | 100.0 |  |  |

| **For how many years have you been active as a manuscript peer reviewer?** | | | | | |
| --- | --- | --- | --- | --- | --- |
|  | | Frequency | Percent | Valid Percent | Cumulative Percent |
| Valid | < 1 year | 11 | 6.4 | 6.5 | 6.5 |
|  | 1-5 years | 59 | 34.3 | 34.9 | 41.4 |
|  | 6-10 years | 43 | 25.0 | 25.4 | 66.9 |
|  | 11-15 years | 15 | 8.7 | 8.9 | 75.7 |
|  | 16-20 years | 13 | 7.6 | 7.7 | 83.4 |
|  | 21 + years | 28 | 16.3 | 16.6 | 100.0 |
|  | Total | 169 | 98.3 | 100.0 |  |
| Missing | System | 3 | 1.7 |  |  |
| Total | | 172 | 100.0 |  |  |

| **How many peer reviewed articles have you published to date?** | | | | | |
| --- | --- | --- | --- | --- | --- |
|  | | Frequency | Percent | Valid Percent | Cumulative Percent |
| Valid | < 2 | 5 | 2.9 | 2.9 | 2.9 |
|  | 3-5 | 16 | 9.3 | 9.3 | 12.2 |
|  | 6-10 | 22 | 12.8 | 12.8 | 25.0 |
|  | 11-20 | 23 | 13.4 | 13.4 | 38.4 |
|  | 21-50 | 36 | 20.9 | 20.9 | 59.3 |
|  | 51+ | 70 | 40.7 | 40.7 | 100.0 |
|  | Total | 172 | 100.0 | 100.0 |  |

| **Have you completed any formal training in peer review?** | | | | | |
| --- | --- | --- | --- | --- | --- |
|  | | Frequency | Percent | Valid Percent | Cumulative Percent |
| Valid | Yes | 26 | 15.1 | 15.2 | 15.2 |
|  | No | 144 | 83.7 | 84.2 | 99.4 |
|  | Unsure | 1 | .6 | .6 | 100.0 |
|  | Total | 171 | 99.4 | 100.0 |  |
| Missing | System | 1 | .6 |  |  |
| Total | | 172 | 100.0 |  |  |

| **What type of formal training received?** | | | | |
| --- | --- | --- | --- | --- |
|  | | Responses | | Percent of Cases |
|  |  | N | Percent |  |
| What type of formal training received? | Online lecture | 10 | 16.4% | 37.0% |
|  | Online course (at least 6 sessions) | 10 | 16.4% | 37.0% |
|  | In-person lecture | 12 | 19.7% | 44.4% |
|  | In-person half day workshop | 2 | 3.3% | 7.4% |
|  | In-person full day workshop | 7 | 11.5% | 25.9% |
|  | Shawdowing a mentor/ghost-writing | 4 | 6.6% | 14.8% |
|  | Self-selected reading material | 7 | 11.5% | 25.9% |
|  | Online resource/modules | 8 | 13.1% | 29.6% |
|  | Other | 1 | 1.6% | 3.7% |
| Total | | 61 | 100.0% | 225.9% |

| **Who provided the training you received?** | | | | |
| --- | --- | --- | --- | --- |
|  | | Responses | | Percent of Cases |
|  |  | N | Percent |  |
| Who provided the traning you received? | A journal | 4 | 12.1% | 14.8% |
|  | A publisher | 6 | 18.2% | 22.2% |
|  | A university/college | 18 | 54.5% | 66.7% |
|  | Private company | 2 | 6.1% | 7.4% |
|  | Unsure/Don't know | 2 | 6.1% | 7.4% |
|  | Other | 1 | 3.0% | 3.7% |
| Total | | 33 | 100.0% | 122.2% |

| **When did you receive the training?** | | | | | |
| --- | --- | --- | --- | --- | --- |
|  | | Frequency | Percent | Valid Percent | Cumulative Percent |
| Valid | ≤1 year ago | 4 | 2.3 | 14.8 | 14.8 |
|  | 2 years ago | 4 | 2.3 | 14.8 | 29.6 |
|  | 3 years ago | 6 | 3.5 | 22.2 | 51.9 |
|  | 4 years ago | 2 | 1.2 | 7.4 | 59.3 |
|  | ≥5 years ago | 11 | 6.4 | 40.7 | 100.0 |
|  | Total | 27 | 15.7 | 100.0 |  |
| Missing | System | 145 | 84.3 |  |  |
| Total | | 172 | 100.0 |  |  |

| **Does the primary institution you are affiliated with offer formal training for peer review?** | | | | | |
| --- | --- | --- | --- | --- | --- |
|  | | Frequency | Percent | Valid Percent | Cumulative Percent |
| Valid | Yes and I have completed it | 10 | 5.8 | 5.8 | 5.8 |
|  | Yes but I have not completed it | 5 | 2.9 | 2.9 | 8.8 |
|  | No | 108 | 62.8 | 63.2 | 71.9 |
|  | Unsure/don't know | 48 | 27.9 | 28.1 | 100.0 |
|  | Total | 171 | 99.4 | 100.0 |  |
| Missing | System | 1 | .6 |  |  |
| Total | | 172 | 100.0 |  |  |

| **Type of training your institution offers** | | | | |
| --- | --- | --- | --- | --- |
|  | | Responses | | Percent of Cases |
|  |  | N | Percent |  |
| Type of training your institution offers | Online lecture | 6 | 20.7% | 37.5% |
|  | Online course (at least 6 sessions) | 2 | 6.9% | 12.5% |
|  | In-person lecture | 3 | 10.3% | 18.8% |
|  | In-person half day workshop | 3 | 10.3% | 18.8% |
|  | In-person full day workshop | 5 | 17.2% | 31.3% |
|  | Shadowing a mentor/ghost-writing | 2 | 6.9% | 12.5% |
|  | Self-selected reading material | 2 | 6.9% | 12.5% |
|  | Online resource/modules | 4 | 13.8% | 25.0% |
|  | Unsure/Don't know | 1 | 3.4% | 6.3% |
|  | Other | 1 | 3.4% | 6.3% |
| Total | | 29 | 100.0% | 181.3% |

| **The first time you did a peer review, how well prepared did you feel you were?** | | | | | |
| --- | --- | --- | --- | --- | --- |
|  | | Frequency | Percent | Valid Percent | Cumulative Percent |
| Valid | Very unprepared | 18 | 10.5 | 10.8 | 10.8 |
|  | Unprepared | 40 | 23.3 | 24.1 | 34.9 |
|  | Slightly unprepared | 30 | 17.4 | 18.1 | 53.0 |
|  | Neutral/Unsure | 11 | 6.4 | 6.6 | 59.6 |
|  | Slightly prepared | 34 | 19.8 | 20.5 | 80.1 |
|  | Prepared | 25 | 14.5 | 15.1 | 95.2 |
|  | Very prepared | 8 | 4.7 | 4.8 | 100.0 |
|  | Total | 166 | 96.5 | 100.0 |  |
| Missing | 0 | 6 | 3.5 |  |  |
| Total | | 172 | 100.0 |  |  |

| **How well prepared do you feel you are to act as a peer reviewer currently?** | | | | | |
| --- | --- | --- | --- | --- | --- |
|  | | Frequency | Percent | Valid Percent | Cumulative Percent |
| Valid | Very unprepared | 2 | 1.2 | 1.2 | 1.2 |
|  | Unprepared | 5 | 2.9 | 3.0 | 4.2 |
|  | Slightly unprepared | 5 | 2.9 | 3.0 | 7.2 |
|  | Neutral/Unsure | 5 | 2.9 | 3.0 | 10.2 |
|  | Slightly prepared | 25 | 14.5 | 15.0 | 25.1 |
|  | Prepared | 66 | 38.4 | 39.5 | 64.7 |
|  | Very prepared | 59 | 34.3 | 35.3 | 100.0 |
|  | Total | 167 | 97.1 | 100.0 |  |
| Missing | 0 | 2 | 1.2 |  |  |
|  | System | 3 | 1.7 |  |  |
|  | Total | 5 | 2.9 |  |  |
| Total | | 172 | 100.0 |  |  |

| **Skills you can improve in peer review** | | | | |
| --- | --- | --- | --- | --- |
|  | | Responses | | Percent of Cases |
|  |  | N | Percent |  |
| Skills you can improve in peer review | None of the above | 14 | 3.0% | 8.1% |
|  | Time Management | 54 | 11.5% | 31.4% |
|  | Structing a review | 67 | 14.3% | 39.0% |
|  | Critical appraisal of theory | 60 | 12.8% | 34.9% |
|  | Critical appraisal of methods | 68 | 14.5% | 39.5% |
|  | Critical appraisal of statistics | 94 | 20.0% | 54.7% |
|  | Understanding of peer reviewer expectations | 59 | 12.6% | 34.3% |
|  | If asked by the journal, making a ‘decision’ on whether to accept/revise/reject a paper | 44 | 9.4% | 25.6% |
|  | Other (please specify) | 10 | 2.1% | 5.8% |
| Total | | 470 | 100.0% | 273.3% |

| **Peer review is important for ensuring the quality and integrity of scholarly communication** | | | | | |
| --- | --- | --- | --- | --- | --- |
|  | | Frequency | Percent | Valid Percent | Cumulative Percent |
| Valid | Strongly disagree | 3 | 1.7 | 1.8 | 1.8 |
|  | Neutral/Unsure | 4 | 2.3 | 2.4 | 4.1 |
|  | Slightly agree | 15 | 8.7 | 8.8 | 12.9 |
|  | Agree | 54 | 31.4 | 31.8 | 44.7 |
|  | Strongly agree | 94 | 54.7 | 55.3 | 100.0 |
|  | Total | 170 | 98.8 | 100.0 |  |
| Missing | 0 | 1 | .6 |  |  |
|  | System | 1 | .6 |  |  |
|  | Total | 2 | 1.2 |  |  |
| Total | | 172 | 100.0 |  |  |

| **My experience acting as a peer reviewer has been positive** | | | | | |
| --- | --- | --- | --- | --- | --- |
|  | | Frequency | Percent | Valid Percent | Cumulative Percent |
| Valid | Disagree | 3 | 1.7 | 1.8 | 1.8 |
|  | Slightly disagree | 1 | .6 | .6 | 2.4 |
|  | Neutral/Unsure | 13 | 7.6 | 7.9 | 10.4 |
|  | Slightly agree | 31 | 18.0 | 18.9 | 29.3 |
|  | Agree | 72 | 41.9 | 43.9 | 73.2 |
|  | Strongly agree | 44 | 25.6 | 26.8 | 100.0 |
|  | Total | 164 | 95.3 | 100.0 |  |
| Missing | 0 | 3 | 1.7 |  |  |
|  | System | 5 | 2.9 |  |  |
|  | Total | 8 | 4.7 |  |  |
| Total | | 172 | 100.0 |  |  |

| **My experience receiving peer review has been positive** | | | | | |
| --- | --- | --- | --- | --- | --- |
|  | | Frequency | Percent | Valid Percent | Cumulative Percent |
| Valid | Strongly disagree | 2 | 1.2 | 1.2 | 1.2 |
|  | Disagree | 8 | 4.7 | 4.8 | 6.0 |
|  | Slightly disagree | 9 | 5.2 | 5.4 | 11.4 |
|  | Neutral/Unsure | 17 | 9.9 | 10.2 | 21.6 |
|  | Slightly agree | 32 | 18.6 | 19.2 | 40.7 |
|  | Agree | 84 | 48.8 | 50.3 | 91.0 |
|  | Strongly agree | 15 | 8.7 | 9.0 | 100.0 |
|  | Total | 167 | 97.1 | 100.0 |  |
| Missing | 0 | 1 | .6 |  |  |
|  | System | 4 | 2.3 |  |  |
|  | Total | 5 | 2.9 |  |  |
| Total | | 172 | 100.0 |  |  |

| **In general, there is a lack of knowledge and understanding for how to properly conduct peer review** | | | | | |
| --- | --- | --- | --- | --- | --- |
|  | | Frequency | Percent | Valid Percent | Cumulative Percent |
| Valid | Strongly disagree | 2 | 1.2 | 1.2 | 1.2 |
|  | Disagree | 14 | 8.1 | 8.3 | 9.5 |
|  | Slightly disagree | 12 | 7.0 | 7.1 | 16.7 |
|  | Neutral/Unsure | 21 | 12.2 | 12.5 | 29.2 |
|  | Slightly agree | 43 | 25.0 | 25.6 | 54.8 |
|  | Agree | 54 | 31.4 | 32.1 | 86.9 |
|  | Strongly agree | 22 | 12.8 | 13.1 | 100.0 |
|  | Total | 168 | 97.7 | 100.0 |  |
| Missing | System | 4 | 2.3 |  |  |
| Total | | 172 | 100.0 |  |  |

| **Peer reviewers should receive formal training in peer review prior to completing peer review assignments for journals** | | | | | |
| --- | --- | --- | --- | --- | --- |
|  | | Frequency | Percent | Valid Percent | Cumulative Percent |
| Valid | Strongly disagree | 4 | 2.3 | 2.4 | 2.4 |
|  | Disagree | 9 | 5.2 | 5.3 | 7.7 |
|  | Slightly disagree | 8 | 4.7 | 4.7 | 12.4 |
|  | Neutral/Unsure | 20 | 11.6 | 11.8 | 24.3 |
|  | Slightly agree | 29 | 16.9 | 17.2 | 41.4 |
|  | Agree | 58 | 33.7 | 34.3 | 75.7 |
|  | Strongly agree | 41 | 23.8 | 24.3 | 100.0 |
|  | Total | 169 | 98.3 | 100.0 |  |
| Missing | System | 3 | 1.7 |  |  |
| Total | | 172 | 100.0 |  |  |

| **My institution values that I contribute to my research field by acting as a peer reviewer** | | | | | |
| --- | --- | --- | --- | --- | --- |
|  | | Frequency | Percent | Valid Percent | Cumulative Percent |
| Valid | Strongly disagree | 23 | 13.4 | 13.9 | 13.9 |
|  | Disagree | 30 | 17.4 | 18.1 | 31.9 |
|  | Slightly disagree | 6 | 3.5 | 3.6 | 35.5 |
|  | Neutral/Unsure | 36 | 20.9 | 21.7 | 57.2 |
|  | Slightly agree | 20 | 11.6 | 12.0 | 69.3 |
|  | Agree | 31 | 18.0 | 18.7 | 88.0 |
|  | Strongly agree | 20 | 11.6 | 12.0 | 100.0 |
|  | Total | 166 | 96.5 | 100.0 |  |
| Missing | 0 | 1 | .6 |  |  |
|  | System | 5 | 2.9 |  |  |
|  | Total | 6 | 3.5 |  |  |
| Total | | 172 | 100.0 |  |  |

| **My institution values that I contribute to my research field by acting as a peer reviewer** | | | | | |
| --- | --- | --- | --- | --- | --- |
|  | | Frequency | Percent | Valid Percent | Cumulative Percent |
| Valid | Strongly disagree | 23 | 13.4 | 13.9 | 13.9 |
|  | Disagree | 30 | 17.4 | 18.1 | 31.9 |
|  | Slightly disagree | 6 | 3.5 | 3.6 | 35.5 |
|  | Neutral/Unsure | 36 | 20.9 | 21.7 | 57.2 |
|  | Slightly agree | 20 | 11.6 | 12.0 | 69.3 |
|  | Agree | 30 | 17.4 | 18.1 | 87.3 |
|  | Strongly agree | 21 | 12.2 | 12.7 | 100.0 |
|  | Total | 166 | 96.5 | 100.0 |  |
| Missing | 0 | 1 | .6 |  |  |
|  | System | 5 | 2.9 |  |  |
|  | Total | 6 | 3.5 |  |  |
| Total | | 172 | 100.0 |  |  |

| **There are appropriate incentives in place to motivate me to engage in peer review** | | | | | |
| --- | --- | --- | --- | --- | --- |
|  | | Frequency | Percent | Valid Percent | Cumulative Percent |
| Valid | Strongly disagree | 41 | 23.8 | 24.6 | 24.6 |
|  | Disagree | 45 | 26.2 | 26.9 | 51.5 |
|  | Slightly disagree | 22 | 12.8 | 13.2 | 64.7 |
|  | Neutral/Unsure | 22 | 12.8 | 13.2 | 77.8 |
|  | Slightly agree | 18 | 10.5 | 10.8 | 88.6 |
|  | Agree | 14 | 8.1 | 8.4 | 97.0 |
|  | Strongly agree | 5 | 2.9 | 3.0 | 100.0 |
|  | Total | 167 | 97.1 | 100.0 |  |
| Missing | 0 | 2 | 1.2 |  |  |
|  | System | 3 | 1.7 |  |  |
|  | Total | 5 | 2.9 |  |  |
| Total | | 172 | 100.0 |  |  |

| **Topics to be covered in PR training** | | | | |
| --- | --- | --- | --- | --- |
|  | | Responses | | Percent of Cases |
|  |  | N | Percent |  |
| Topics to be covered in PR training | Research question | 28 | 17.5% | 17.5% |
|  | Abstract | 1 | 0.6% | 0.6% |
|  | Study design / methodology | 35 | 21.9% | 21.9% |
|  | Statistics | 33 | 20.6% | 20.6% |
|  | References | 2 | 1.3% | 1.3% |
|  | Discussion | 3 | 1.9% | 1.9% |
|  | Study limitations | 2 | 1.3% | 1.3% |
|  | Supplementary reporting | 4 | 2.5% | 2.5% |
|  | Concerns of publication ethics (ex. plagiarism, conflicts of interest, misconduct) | 11 | 6.9% | 6.9% |
|  | How to construct a peer review | 41 | 25.6% | 25.6% |
| Total | | 160 | 100.0% | 100.0% |

| **Who is best to offer PR training?** | | | | |
| --- | --- | --- | --- | --- |
|  | | Responses | | Percent of Cases |
|  |  | N | Percent |  |
| Who is best to offer PR training? | University / college | 59 | 36.6% | 36.6% |
|  | Membership societies | 13 | 8.1% | 8.1% |
|  | Scholarly publishers or journals | 65 | 40.4% | 40.4% |
|  | Independent course providers | 6 | 3.7% | 3.7% |
|  | It doesn't matter | 14 | 8.7% | 8.7% |
|  | Peer review training is unnecessary | 4 | 2.5% | 2.5% |
| Total | | 161 | 100.0% | 100.0% |

| **Who should fund PR training?** | | | | |
| --- | --- | --- | --- | --- |
|  | | Responses | | Percent of Cases |
|  |  | N | Percent |  |
| Who should fund PR training? | You (the person being trained) | 12 | 7.3% | 7.3% |
|  | Your primary research institution | 48 | 29.3% | 29.3% |
|  | Scholarly publishers | 79 | 48.2% | 48.2% |
|  | Funders | 10 | 6.1% | 6.1% |
|  | Nobody | 12 | 7.3% | 7.3% |
|  | It does not matter | 3 | 1.8% | 1.8% |
| Total | | 164 | 100.0% | 100.0% |

| **You (the person being trained)** | | | | | |
| --- | --- | --- | --- | --- | --- |
|  | | Frequency | Percent | Valid Percent | Cumulative Percent |
| Valid | 1 | 12 | 7.0 | 9.3 | 9.3 |
|  | 2 | 13 | 7.6 | 10.1 | 19.4 |
|  | 3 | 19 | 11.0 | 14.7 | 34.1 |
|  | 4 | 34 | 19.8 | 26.4 | 60.5 |
|  | 5 | 19 | 11.0 | 14.7 | 75.2 |
|  | 6 | 32 | 18.6 | 24.8 | 100.0 |
|  | Total | 129 | 75.0 | 100.0 |  |
| Missing | System | 43 | 25.0 |  |  |
| Total | | 172 | 100.0 |  |  |

| **Your primary research institution** | | | | | |
| --- | --- | --- | --- | --- | --- |
|  | | Frequency | Percent | Valid Percent | Cumulative Percent |
| Valid | 1 | 48 | 27.9 | 32.7 | 32.7 |
|  | 2 | 45 | 26.2 | 30.6 | 63.3 |
|  | 3 | 38 | 22.1 | 25.9 | 89.1 |
|  | 4 | 8 | 4.7 | 5.4 | 94.6 |
|  | 5 | 4 | 2.3 | 2.7 | 97.3 |
|  | 6 | 4 | 2.3 | 2.7 | 100.0 |
|  | Total | 147 | 85.5 | 100.0 |  |
| Missing | System | 25 | 14.5 |  |  |
| Total | | 172 | 100.0 |  |  |

| **Scholarly publishers** | | | | | |
| --- | --- | --- | --- | --- | --- |
|  | | Frequency | Percent | Valid Percent | Cumulative Percent |
| Valid | 1 | 79 | 45.9 | 49.1 | 49.1 |
|  | 2 | 46 | 26.7 | 28.6 | 77.6 |
|  | 3 | 22 | 12.8 | 13.7 | 91.3 |
|  | 4 | 10 | 5.8 | 6.2 | 97.5 |
|  | 5 | 1 | .6 | .6 | 98.1 |
|  | 6 | 3 | 1.7 | 1.9 | 100.0 |
|  | Total | 161 | 93.6 | 100.0 |  |
| Missing | System | 11 | 6.4 |  |  |
| Total | | 172 | 100.0 |  |  |

| **Funders** | | | | | |
| --- | --- | --- | --- | --- | --- |
|  | | Frequency | Percent | Valid Percent | Cumulative Percent |
| Valid | 1 | 10 | 5.8 | 7.1 | 7.1 |
|  | 2 | 36 | 20.9 | 25.5 | 32.6 |
|  | 3 | 51 | 29.7 | 36.2 | 68.8 |
|  | 4 | 36 | 20.9 | 25.5 | 94.3 |
|  | 5 | 5 | 2.9 | 3.5 | 97.9 |
|  | 6 | 3 | 1.7 | 2.1 | 100.0 |
|  | Total | 141 | 82.0 | 100.0 |  |
| Missing | System | 31 | 18.0 |  |  |
| Total | | 172 | 100.0 |  |  |

| **Nobody** | | | | | |
| --- | --- | --- | --- | --- | --- |
|  | | Frequency | Percent | Valid Percent | Cumulative Percent |
| Valid | 1 | 12 | 7.0 | 9.8 | 9.8 |
|  | 2 | 2 | 1.2 | 1.6 | 11.4 |
|  | 3 | 3 | 1.7 | 2.4 | 13.8 |
|  | 4 | 21 | 12.2 | 17.1 | 30.9 |
|  | 5 | 58 | 33.7 | 47.2 | 78.0 |
|  | 6 | 27 | 15.7 | 22.0 | 100.0 |
|  | Total | 123 | 71.5 | 100.0 |  |
| Missing | System | 49 | 28.5 |  |  |
| Total | | 172 | 100.0 |  |  |

| **It does not matter** | | | | | |
| --- | --- | --- | --- | --- | --- |
|  | | Frequency | Percent | Valid Percent | Cumulative Percent |
| Valid | 1 | 3 | 1.7 | 2.4 | 2.4 |
|  | 2 | 6 | 3.5 | 4.8 | 7.3 |
|  | 3 | 4 | 2.3 | 3.2 | 10.5 |
|  | 4 | 19 | 11.0 | 15.3 | 25.8 |
|  | 5 | 35 | 20.3 | 28.2 | 54.0 |
|  | 6 | 57 | 33.1 | 46.0 | 100.0 |
|  | Total | 124 | 72.1 | 100.0 |  |
| Missing | System | 48 | 27.9 |  |  |
| Total | | 172 | 100.0 |  |  |

| **What is best method for PR training delivery?** | | | | |
| --- | --- | --- | --- | --- |
|  | | Responses | | Percent of Cases |
|  |  | N | Percent |  |
| What is best method for PR trng delivery | Online lecture | 53 | 33.5% | 33.5% |
|  | Online course (at least 6 sessions) | 37 | 23.4% | 23.4% |
|  | In-person lecture | 7 | 4.4% | 4.4% |
|  | In-person half day workshop | 8 | 5.1% | 5.1% |
|  | In-person full day workshop | 15 | 9.5% | 9.5% |
|  | Shadowing a mentor/ghost-writing | 17 | 10.8% | 10.8% |
|  | Self-selected reading material | 3 | 1.9% | 1.9% |
|  | Online resources/modules | 18 | 11.4% | 11.4% |
| Total | | 158 | 100.0% | 100.0% |

| **Do you operate or work or volunteer for a journal that publishes peer reviewed articles?** | | | | | |
| --- | --- | --- | --- | --- | --- |
|  | | Frequency | Percent | Valid Percent | Cumulative Percent |
| Valid | Yes, continue to this section | 79 | 45.9 | 46.7 | 46.7 |
|  | No, skip this section | 90 | 52.3 | 53.3 | 100.0 |
|  | Total | 169 | 98.3 | 100.0 |  |
| Missing | System | 3 | 1.7 |  |  |
| Total | | 172 | 100.0 |  |  |

| **What is your role at the journal?** | | | | | |
| --- | --- | --- | --- | --- | --- |
|  | | Frequency | Percent | Valid Percent | Cumulative Percent |
| Valid | Other (please specify) | 28 | 16.3 | 33.7 | 33.7 |
|  | Editor in chief | 7 | 4.1 | 8.4 | 42.2 |
|  | Editorial board member | 48 | 27.9 | 57.8 | 100.0 |
|  | Total | 83 | 48.3 | 100.0 |  |
| Missing | System | 89 | 51.7 |  |  |
| Total | | 172 | 100.0 |  |  |

| **Does the journal have explicit eligibility criteria for selecting peer reviewers?** | | | | | |
| --- | --- | --- | --- | --- | --- |
|  | | Frequency | Percent | Valid Percent | Cumulative Percent |
| Valid | Yes | 31 | 18.0 | 38.3 | 38.3 |
|  | No | 27 | 15.7 | 33.3 | 71.6 |
|  | Unsure/Don't know | 23 | 13.4 | 28.4 | 100.0 |
|  | Total | 81 | 47.1 | 100.0 |  |
| Missing | System | 91 | 52.9 |  |  |
| Total | | 172 | 100.0 |  |  |

| **Does the journal require any explicit training prior to allowing peer reviewers to assess a manuscript?** | | | | | |
| --- | --- | --- | --- | --- | --- |
|  | | Frequency | Percent | Valid Percent | Cumulative Percent |
| Valid | Yes, internally provided training | 8 | 4.7 | 10.0 | 10.0 |
|  | Yes, externally provided training | 2 | 1.2 | 2.5 | 12.5 |
|  | No | 55 | 32.0 | 68.8 | 81.3 |
|  | Unsure/Don't know | 15 | 8.7 | 18.8 | 100.0 |
|  | Total | 80 | 46.5 | 100.0 |  |
| Missing | System | 92 | 53.5 |  |  |
| Total | | 172 | 100.0 |  |  |

| **What type of PR training be required?** | | | | |
| --- | --- | --- | --- | --- |
|  | | Responses | | Percent of Cases |
|  |  | N | Percent |  |
| What type of PR trng be required? | Online lecture | 6 | 22.2% | 54.5% |
|  | Online course (at least 6 sessions) | 6 | 22.2% | 54.5% |
|  | In-person lecture | 3 | 11.1% | 27.3% |
|  | In-person halfday workshop | 1 | 3.7% | 9.1% |
|  | In-person fullday workshop | 3 | 11.1% | 27.3% |
|  | Shadowing a mentor/ghost-writing | 2 | 7.4% | 18.2% |
|  | Self-selected reading material | 1 | 3.7% | 9.1% |
|  | Online resource/modules | 4 | 14.8% | 36.4% |
|  | Other (please specify) | 1 | 3.7% | 9.1% |
| Total | | 27 | 100.0% | 245.5% |

| **How many hours of peer review training does the journal require before allowing reviewers assess a manuscript?** | | | | | |
| --- | --- | --- | --- | --- | --- |
|  | | Frequency | Percent | Valid Percent | Cumulative Percent |
| Valid | Other (please specify) | 1 | .6 | 9.1 | 9.1 |
|  | No set amount of training | 3 | 1.7 | 27.3 | 36.4 |
|  | 1-5 hours | 2 | 1.2 | 18.2 | 54.5 |
|  | 6-10 hours | 3 | 1.7 | 27.3 | 81.8 |
|  | 15-20 hours | 1 | .6 | 9.1 | 90.9 |
|  | 20 hours + | 1 | .6 | 9.1 | 100.0 |
|  | Total | 11 | 6.4 | 100.0 |  |
| Missing | System | 161 | 93.6 |  |  |
| Total | | 172 | 100.0 |  |  |

| **Does the journal explicitly assess peer review reports of new peer reviewers?** | | | | | |
| --- | --- | --- | --- | --- | --- |
|  | | Frequency | Percent | Valid Percent | Cumulative Percent |
| Valid | Yes (please specify how) | 8 | 4.7 | 10.0 | 10.0 |
|  | No | 21 | 12.2 | 26.3 | 36.3 |
|  | Unsure/Don't know | 51 | 29.7 | 63.7 | 100.0 |
|  | Total | 80 | 46.5 | 100.0 |  |
| Missing | System | 92 | 53.5 |  |  |
| Total | | 172 | 100.0 |  |  |

| **Does the journal have a database of peer reviewers?** | | | | | |
| --- | --- | --- | --- | --- | --- |
|  | | Frequency | Percent | Valid Percent | Cumulative Percent |
| Valid | Yes, less than 50 reviewers | 13 | 7.6 | 16.3 | 16.3 |
|  | Yes, more than 50 reviewers | 44 | 25.6 | 55.0 | 71.3 |
|  | No | 3 | 1.7 | 3.8 | 75.0 |
|  | Unsure/don't know | 20 | 11.6 | 25.0 | 100.0 |
|  | Total | 80 | 46.5 | 100.0 |  |
| Missing | System | 92 | 53.5 |  |  |
| Total | | 172 | 100.0 |  |  |

| **Does the journal explicitly provide reporting guidelines to reviewers as part of the peer review assessment process?** | | | | | |
| --- | --- | --- | --- | --- | --- |
|  | | Frequency | Percent | Valid Percent | Cumulative Percent |
| Valid | Yes | 51 | 29.7 | 64.6 | 64.6 |
|  | No | 21 | 12.2 | 26.6 | 91.1 |
|  | Unsure/Don't know | 7 | 4.1 | 8.9 | 100.0 |
|  | Total | 79 | 45.9 | 100.0 |  |
| Missing | System | 93 | 54.1 |  |  |
| Total | | 172 | 100.0 |  |  |
