## Supplementary figures and images for "Knowledge and motivations of training in peer review: an international cross-sectional survey"

### Email flowchart

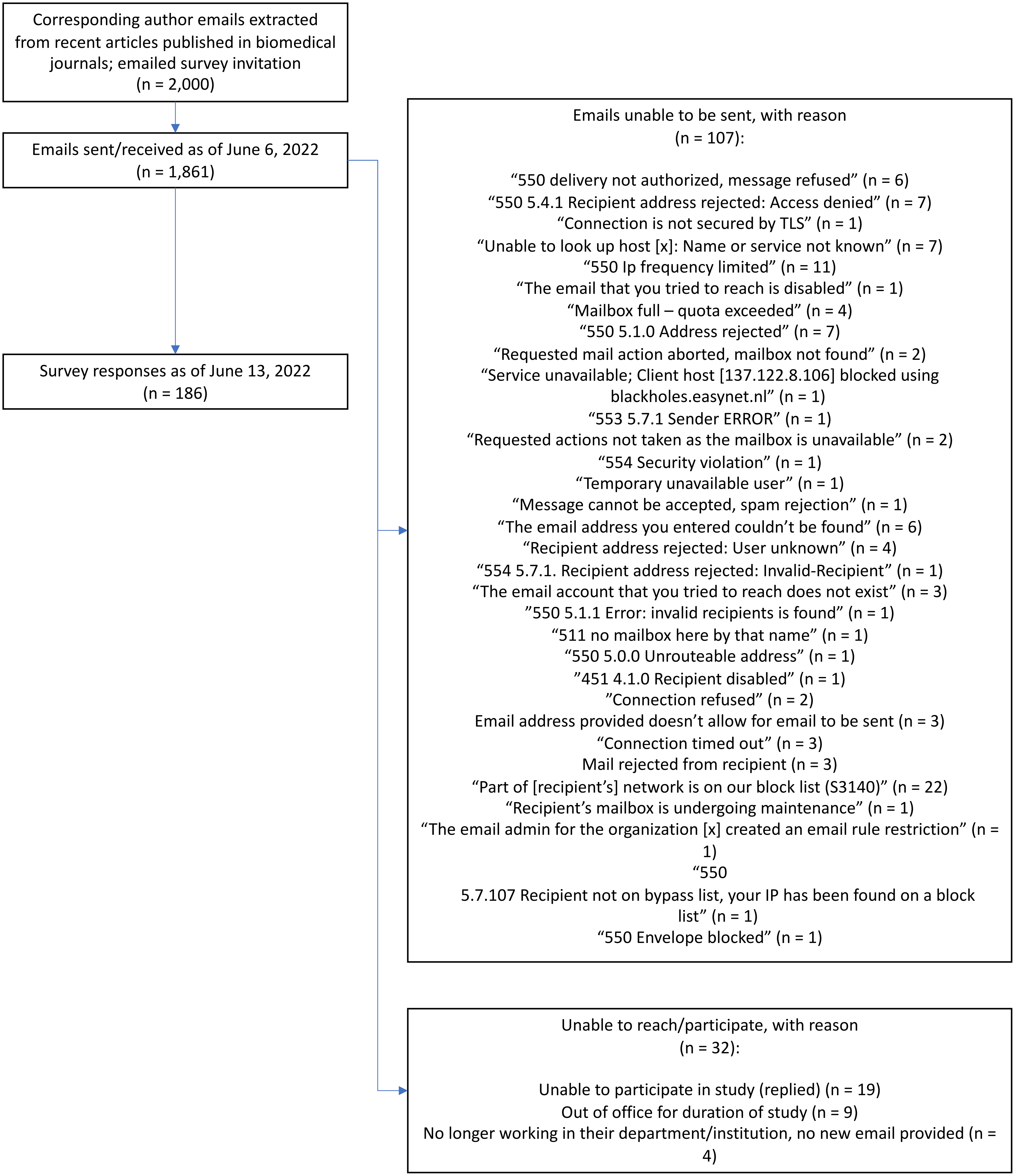
